# Phylogenetic analysis of kinetoplast DNA: kDNA of *Leishmania tropica* in Thi-Qar province, Iraq

**DOI:** 10.1101/2020.10.28.20220228

**Authors:** Mohammed Hassan Flaih, Fadhil Abbas Al-Abady, Khwam Reissan Hussein

**Affiliations:** Medical Laboratory Department, Al-Nasiriya Technical Institute, Southern Technical University, Thi-Qar, Iraq; Department of Biology, College of Education for pure Sciences, University of Thi-Qar, Thi-Qar, Iraq

**Keywords:** Cutaneous Leishmaniasis, *Leishmania tropica*, Phylogenetic tree, kDNA sequences

## Abstract

**Introduction:** Cutaneous leishmaniasis (CL) is one of wobbling endemic disease in Iraq, that cause intracellular obligate protistan parasite returned to the genus *Leishmania*. This study is aimed to identify epidemiology of CL, detect the prevalence of *Leishmania tropica* and find the phylogenetic relationship.

**Methodology:** The current study was conducted in the main hospitals of Thi-Qar province-south of Iraq for period from November 2018 to October 2019. Nested-PCR was used to amplify kinetoplast minicircle fragments DNA.

**Results:** It was recorded 247 clinical cases with CL, the infections of males were higher than females, while infection rate appeared gradual reduction with age progress. Furthermore, the most CL infections were as single lesions and occurred in December. The infections of upper limbs were high when compared with other body regions. The molecular diagnosis showed *L. tropica* was more frequently. DNA sequences of *kDNA* gene of *L. tropica* showed confirmative genetic detection of local isolates using NCBI-Blast data and phylogenetic tree analysis after comparison with global recorded isolates. The local *L. tropica* isolates showed genetically closed related to NCBI-Blast *L. tropica* with accession number AB678350.1. Generally, the analysis of kDNA nitrogen bases sequences showed that all of samples were consistent with those recorded at the NCBI.

**Conclusion:** The kDNA minicircle sequences analysis results showed mismatching of the local isolates decrease whenever approached from the Iranian border. In addition, genetic heterogeneity diagnosis is important for detection of therapy, control and epidemiological studies.

## Introduction

Leishmaniasis is a vector-borne disease, that causes intracellular obligate protistan parasite returned to the genus *Leishmania* (1). There are nearly twenty one known *Leishmania* species, that may cause different clinical symptoms in humans (2). Ninety percent from cases of cutaneous leishmaniasis (CL) occur in Afghanistan, Algeria, Brazil, Iran, Peru, Saudi Arabia and Syria (3). *Leishmania tropica* and *L. major* usually cause CL and have same life cycle, but they found in different localities and they have different intermediate and reservoir hosts (4). Although pathogenicity, virulence and clinical symptoms vary among *Leishmania* species, it also depends on sandfly type and the genetic background of host (2). Overall, the infection begins as a small erythema around the site of the sandfly bite, gradually converts to an inflammatory papule then increases in size, after that becomes a nodule not painful. Finally, it appears discoloration scar (5).

The diagnosis of CL depends on clinical case and laboratorial examinations as parasitological, serological and molecular tests (6). However, traditional methods of diagnosis as culture media and smear preparation do not discriminate causative species of disease (7). Molecular examinations are widely used for epidemiological and diagnostic applications for determine the parasite at genotype level (8). There are molecular techniques used in determination of *Leishmania* genus, species or intra-species that depend on different target genes such as miniexon (9), internal transcribed spacer of ribosomal DNA (ITS-rDNA) (10), small subunit ribosomal-RNA region (SSU-rRNA), HSP-70, CPb, triose-phosphate isomerase (tim) (11), 18S-ribosomal RNA (rRNA), Gp63, kinetoplast DNA (kDNA) and cytochrome b (*Cyt b*) (12). However, *Leishmania* parasites have a single branched mitochondrion (kinetoplast), that contains a large-network of kDNA (13, 2). *kDNA* gene has shown highly sensitivity and specificity in detecting *Leishmania* (14).

The population structure and phylogenetic covariance at strain/haplotype level of *Leishmania spp*. may provide route of identifying epidemiological changes that occur leishmaniasis (15). Indeed, the analysis of genetic diversity of *Leishmania spp*. is one of important methods not only for control and diagnosis, but also for relevant epidemiologic and taxonomic relationships (16). kDNA minicircle sequencing or PCR-RFLP has shown difference among strains genetically homogenous with other techniques (17). In short, the kDNA minicircle of *Leishmania* parasite is provide an ideal target in order to the genotyping, because the sequence differences are allowing for correct discrimination between species (18).

Considering to importance of Thi-Qar province as a one of the mixed foci of cutaneous and visceral leishmaniasis. To our knowledge, the molecular characterization and phylogenetic relationship of *L. tropica* isolates have not been studied before in the local area. One of the primary aims of this study is to identify the epidemiology of CL in spatially and timetable epidemic areas. Also, to determine the rate of *L. tropica* prevalence in the local area by amplifying kinetoplast minicircle DNA with nested-PCR, and characterized by kDNA sequencing and phylogenetic tree to examine the intra-species relationships within *L. tropica*.

## Methodology

### Geographical area and patients

Thi-Qar is large province in south of Iraq (Figure 1) and locates at 31°14′N 46°19′E. The total land reaches 12,900 km^2^, the province’s population is nearly 2 million people (19). The field of the study included three hospitals: Al-Hussein Teaching, Al-Suoq and Al-Shatrah, for the period from November 2018 to October 2019. The hospitals have received patients suffering from cutaneous diseases. All medical information was taken from patient records of public health department, Thi-Qar health office, including gender, age, date and location and number of lesions. The samples (80) were collected randomly from patients with CL. The lesions were injected using normal saline (0.2 ml) in lesion edge, then pulled again, after that the fluid was kept in plain tube.

**Figure 1:**
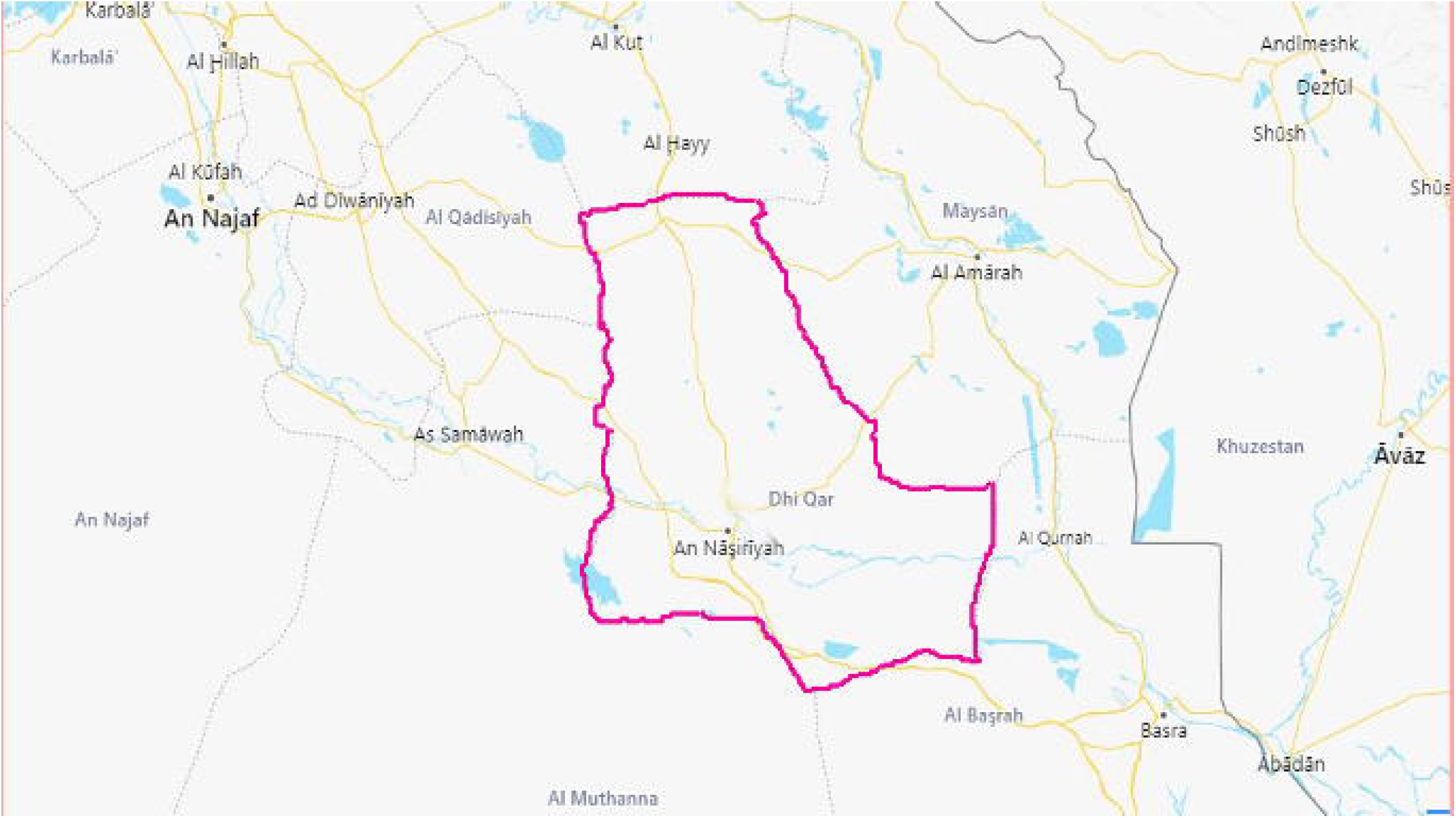
The location and border of Thi-Qar province. https://www.bing.com/maps?q=iraq&search=&form=QBLH#

Ethical approval was received from College of Education for pure Sciences, University of Thi-Qar, Thi-Qar, Iraq.

### Genomic DNA extraction

The gSYAN DNA kit (Geneaid) was used for DNA extraction from lesion fluid, according to protocol of producing company. Extracted DNA was examined using Nanodrop spectrophotometer, then stored at -20°C until used in PCR amplification.

### Nested-PCR amplification

Nested PCR involves two primers with two successive runs. The second primer amplifies a target within the first run product (20). Kinetoplast DNA was amplified for determination of *Leishmania spp*. using Nested-PCR according to the Noyes’s technique (21), which includes two steps. Target DNA was run with external primers: CSB2XF 5’-CGA GTA GCA GAA ACT CCC GTT CA-3’ and CSB1XR 5’-ATT TTT CGC GAT TTT CGC AGA ACG-3’, then first run product undergone to second run with internal specific primers:13Z 5’-ACT GGG GGT TGG TGT AAA ATA G-3’ and LiR 5’-TCG CAG AAC GCC CCT-3’. PCR-master mix was prepared according to the AccuPower® PCR PreMix kit (Bioneer, Korea). Nested PCR primers were provided by Macrogen Company, Korea. The master mix prepared 5μL of genomic DNA, 10 pmol of each external primer and 13 μL from molecular grade water (MGW) and placed in Eppendorf tubes. The thermal conditions of PCR reaction included initial denaturation at 95 □ C for 5 min followed by 30 cycles at 95°C for 30 sec, 55°C for 30 sec and 72°C for one min, and final extension 72°C for 5 min. Nested PCR master mix of second run included 3 μL of first run product, 10 pmol of each internal primer and 15 μL of MGW and placed in Eppendorf tubes. The PCR program was run. The PCR products were passed electrophoresis in agarose gel (1%) with 3μL of ethidium bromide. PCR product (10 μL was added in to each comb well and 5 μl of 100bp ladder in each well. Gel tray was fixed in electrophoresis chamber and filled with 1X TBE buffer. Then electric current passed at 100 volts and 80 AM for 1hr. PCR products were visualized with ultraviolet transilluminator.

### Sequencing and phylogenetic tree

The kDNA samples were visualized on the gel, then purified using a gel purification kit and then sent to Macrogene (Korean Company) for sequence analysis. The kDNA sequences were received then submitted to GenBank in order to obtain accession numbers. These records were compared with deposited accession numbers in GeneBank to find the similarity and mismatching between kDNA sequence of local *L. tropica* isolates and NCBI-Genbank *L. tropica* strains using the BLAST analysis software, (http://blast.ncbi.nlm.nih.gov/Blast.cgi). The kDNA sequences were trimmed by ClustalW alignment analysis by using (MEGA 6.0, multiple alignment analysis tool). Phylogenetic tree was performed with multiple sequence alignment analysis using phylogenetic UPGMA tree type (22).

### Statistical Analysis

In the study, Chi-Square (χ ^2^) test was used in order to analyze the data using SPSS statistical package software V.17. Significant level was set at (P ≤0.05).

## Results

### Epidemiological data

A total of 247 suspected cases with cutaneous leishmaniasis, 80 clinical samples were selected randomly for molecular examination from patients with single or multi-lesion. Generally, 247 patients were divided 164 males and 83 females (Table1). The age group ≤10 years old recorded significantly higher infection rate (p < 0.05) than other age groups. Most of the cases appeared as a single lesion (55.47%) (Table 2). Upper limbs were recorded higher infection rate which reached 18.6% than lower limbs (17.4%) of the total single lesions. There were significant differences at p < 0.05 when compared with other lesion locations. Also, the results showed the infection rate was high in December and observed significant variation when compared with other months.

**Table1:** Age and gender distribution of CL patients

| Age group<br>(years) | Males | Females | Total | <u>Table1: Age and gender</u><br><u>distribution of CL</u><br><u>patients</u> |
| --- | --- | --- | --- | --- |
|  | <u>No (%)</u> | <u>No (%)</u> | <u>No (%)</u> |  |
| □ 10 | 50 (20.24) | 31(12.55) | 81 (32.8) |  |
| 11-20 | 39 (15.79) | 25 (10.12) | 64 (25.9) |  |
| 21-30 | 35 (14.17) | 12 (4.86) | 47 (19.0) |  |
| 31-40 | 25 (10.12) | 8 (3.24) | 33 (13.4) |  |
| >41 | 15 (9.15) | 7 (8.4) | 22 (8.9) |  |
| Total | 164 (66.4) | 83 (33.6) | 247 (100) |  |

**Table 2:**
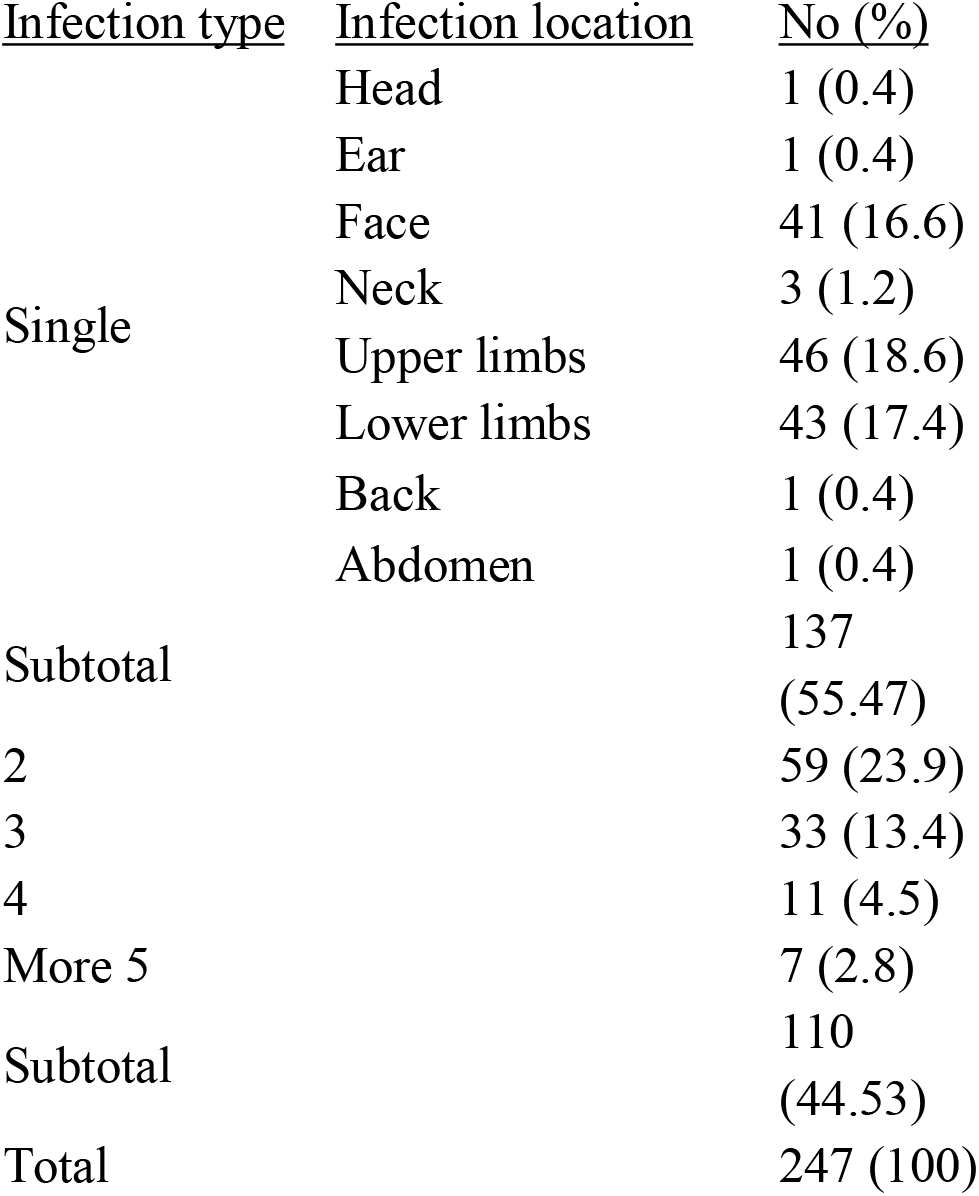
Anatomical location and type of lesion of CL

**Table 3:**
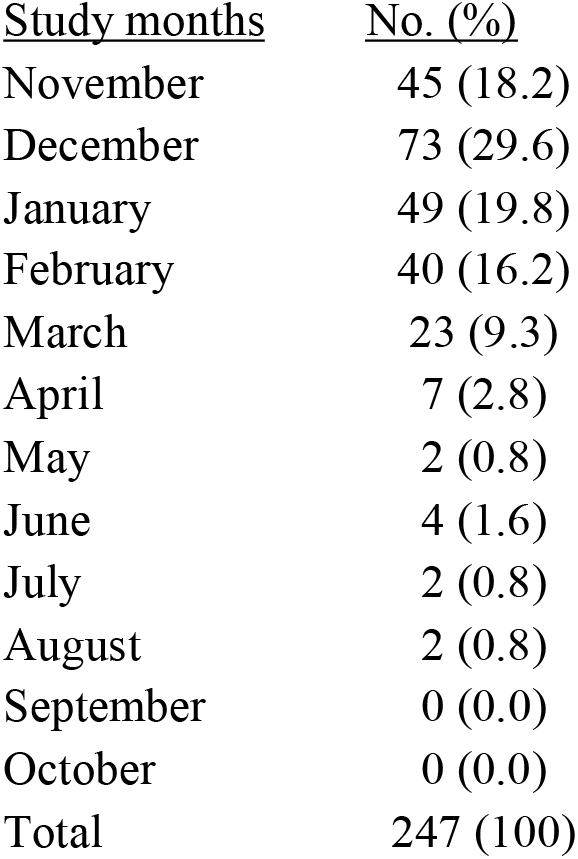
Infection distribution according to study months

| <u>Study months</u> | <u>No. (%)</u> |
| --- | --- |
| November | 45 (18.2) |
| December | 73 (29.6) |
| January | 49 (19.8) |
| February | 40 (16.2) |
| March | 23 (9.3) |
| April | 7 (2.8) |
| May | 2 (0.8) |
| June | 4 (1.6) |
| July | 2 (0.8) |
| August | 2 (0.8) |
| September | 0 (0.0) |
| October | 0 (0.0) |
| Total | 247 (100) |

### Molecular identification

The kDNA was amplified using Nested PCR, then electrophoresed through agarose gel (1%) of minicircle fragments, 750 bp of *L. tropica* (Figure 2). The results showed 65 out of 80 were positive for cutaneous leishmaniasis, and distinguished by two species co-existing in the area. *L. tropica* was the most common species which that comprised 70.77% of the total positive samples, while *L. major* (29.23%) showed in low level.

**Figure 2:**
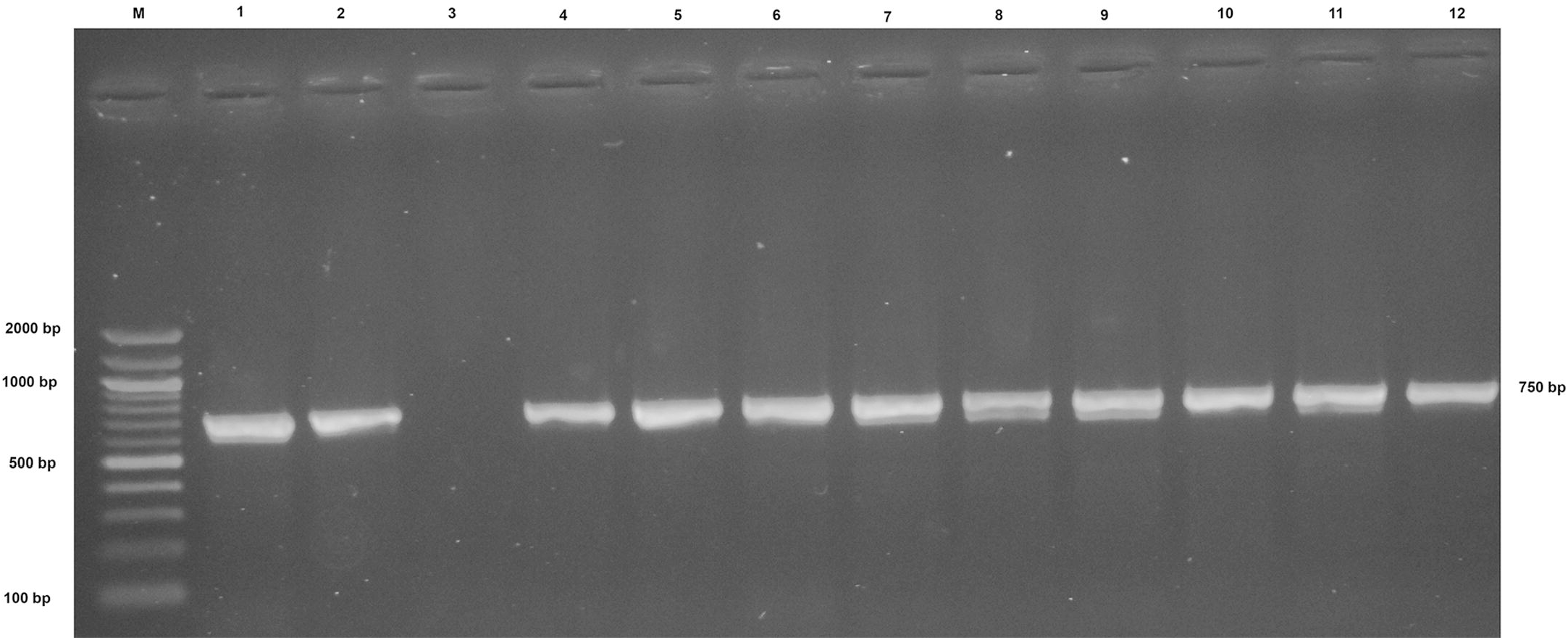
Agarose gel (1%) electrophoresis of *Leishmania* species analyzed using nested-PCR of *kDNA* in CL positive samples. Where M: marker (2000-100bp), lanes 1, 2 and 4-12 *L. tropica* isolated from human skin lesions, while lance 3 negative sample.

### Sequence type and phylogenetic analysis

The nucleotide sequences of kinetoplast minicircle DNA (kDNA) gene of 10 *L. tropica* clinical samples were selected from different places of the study area. However, alignment of sequence type of kDNA (Figure 3) showed in upstream nucleotides different locations and slight nucleotide variation. All samples were consistent with those recorded in the National Center for Biotechnology Information (NCBI) with a ratio of 99% (Table 4). Nucleotides sequences data were submitted to the GenBank-database and recorded in the website with 10 accession numbers MN334661-MN334670. In the current study, phylogenetic tree was designed to determine the extent of genetic affinity between them and the global strains (Figure 4). All the local *L. tropica* isolates showed relatively more similar nitrogen bases sequences and genetically closed related to NCBI-Blast *L. tropica* (AB678350.1) at total genetic change (0.001-0.004).

**Table 4:** NCBI-BLAST Homology Sequence identity between local *L. tropica* isolates and NCBI-Genbank isolates species

| Local *L. tropica* isolates | Genbank submission accession number | NCBI-BLAST Homology Sequence identity |  |  |  |
| --- | --- | --- | --- | --- | --- |
|  |  | NCBI BLAST identity isolate | Country | accession number | Identity (%) |
| IQ.N-No.1 | MN334661 | *L. tropica* | Iran | AB678350.1 | 99.67% |
| IQ.N-No.2 | MN334662 | *L. tropica* | Iran | AB678350.1 | 100% |
| IQ.N-No.3 | MN334663 | *L. tropica* | Iran | AB678350.1 | 100% |
| IQ.N-No.4 | MN334664 | *L. tropica* | Iran | AB678350.1 | 100% |
| IQ.N-No.5 | MN334665 | *L. tropica* | Iran | AB678350.1 | 99.83% |
| IQ.N-No.6 | MN334666 | *L. tropica* | Iran | AB678350.1 | 100% |
| IQ.N-No.7 | MN334667 | *L. tropica* | Iran | AB678350.1 | 99.67% |
| IQ.N-No.8 | MN334668 | *L. tropica* | Iran | AB678350.1 | 100% |
| IQ.N-No.9 | MN334669 | *L. tropica* | Iran | AB678350.1 | 99.67% |
| IQ.N-No.10 | MN334670 | *L. tropica* | Iran | AB678350.1 | 99.67% |

**Figure 3:**
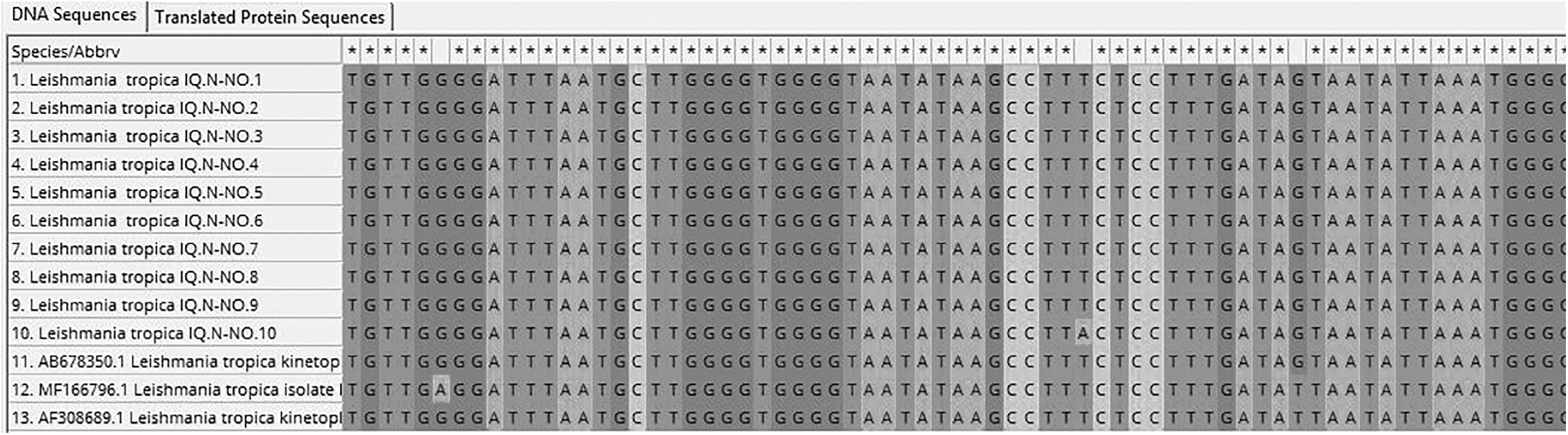
Multiple sequence alignment analysis of partial cutaneous leishmaniasis *kDNA*, non-protein coding region sequence in local *L. tropica* isolates and NCBI-Genbank *L. tropica* isolates based ClustalW alignment analysis using (MEGA 6.0, multiple alignment analysis tool). The multiple alignment analysis showed similarity (*) and mismatching in *kDNA* gene nucleotide sequences.

**Figure 4:**
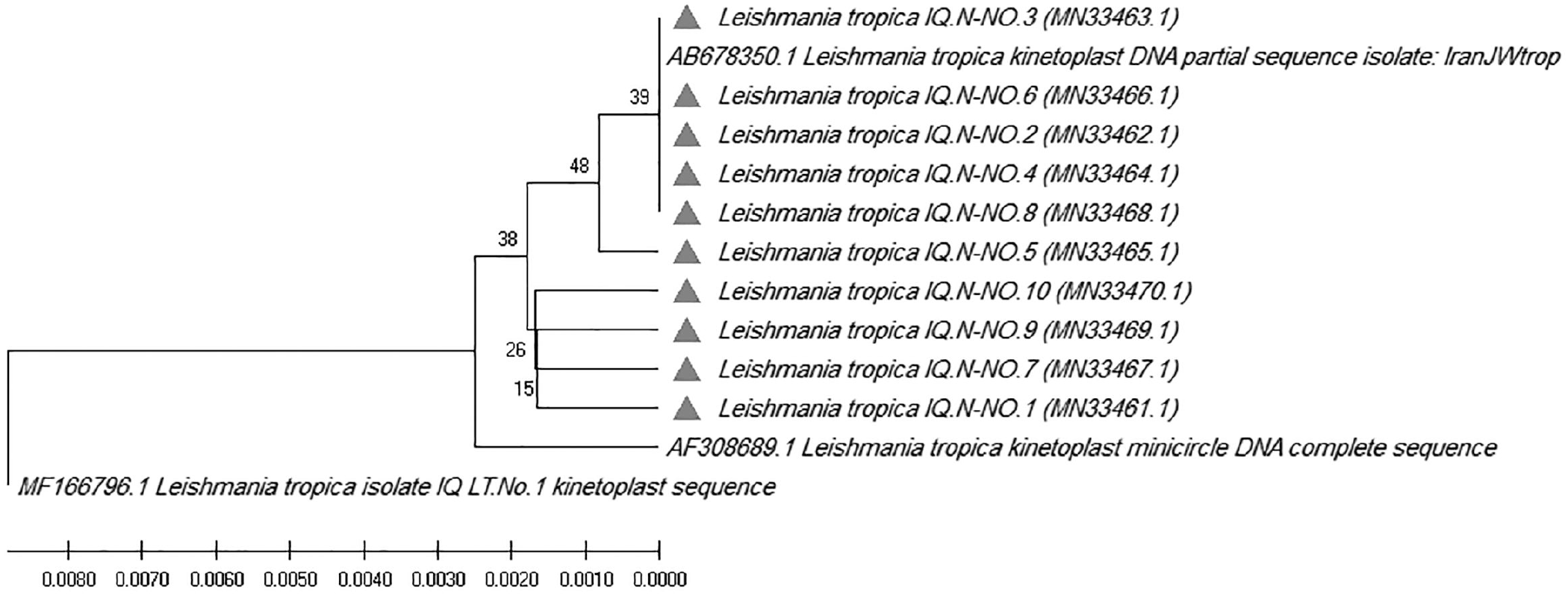
Phylogenetic tree analysis based on the partial sequence of kinetoplast minicircle DNA, non-protein coding region in local *L. tropica* isolates that used for confirmative genetic detection and genetic relationship analysis. The evolutionary distances were computed using phylogenetic UPGMA tree type (MEGA 6.0 version).

## Discussion

Leishmaniasis infections spread in geographically different regions, and more infections occur in sub- or/and tropics of Africa, the Middle East, Southern and Central America and South Europe and Asia (23). The spatial distribution of the disease and identification of the epidemiological data is an essential for future protection (1). However, the traditional diagnosis methods show low sensitivity and difficulty in differentiate the organism at the genus level (24). The parasitological test in most cases is highly sensitive in co-infected cases, but they have low sensitive in a large number of cases (25). In short, PCR methods have highly specific and sensitive to diagnosis of species, with value reaching to 100% (8). Nested and semi-nested PCR are techniques used for discrimination of the species (2).

In the study and at epidemiologic level, there was a significant difference at p < 0.05 between the infection of the males and females. This finding is consistent with Ramezany *et al*. (22). The males who were most exposed to sandfly bite, because they sleep most time uncovered and coming out to play. Also, the results discovered significant differences (p < 0.05) among age groups, ≤10, 11-20 years old recorded the highest infection rates (32.8%, 25.9%) respectively, while the lowest rate was 8.9% within ≥41 years old. This finding is consistent with Ali *et al*. (26). LC decreases with age. The single lesion was 137 (55.46%) cases and multi-lesion recorded 110 (44.53%) cases, while upper limbs were high compared with other single lesions. These findings were identical with Abdellatif *et al*. (27) and Moosazadeh *et al*. (3), but they are not consistent with Al-Hassani (28). The exposed parts and sandflies abundance play a role in the infection incidence. Generally, December recorded the highest rate of the infection when compared with other study months, the result is not agreement with Al-Samarai and Al-Obaidi (29). The findings depended on a review date of patient and no date of appearance of symptoms.

Overall, one of the most common methods used for determination of *Leishmania* infections is extrachromosomal DNA such as kinetoplast DNA [24, 28]. kDNA gene amplification is a suitable to differentiate *Leishmania* species [29,30]. kDNA gene has high sensitivity, kinetoplast includes almost 10,000 circulars and convoluted kDNA minicircles per cell (33). In the current study, most common parasite was *L. topica*, while *L. major* was low incidence. This is consistent with Ramezany *et al*. (24) in south-eastern Iran, and does not agree with Al-Hassani (28) in Al-Qadisiya province, Iraq.

A species identification is important in order epidemiological studies and clinical forms of disease (9). A diagnosis and identification of geographic distribution of *Leishmania* species/strains are required to order appropriate treatment and public health control (8). There are several target genes sequences have been used for the diagnosis of leishmaniasis infections such as kDNA gene (25), rRNA genes (34), miniexon (35), ITS regions (36) and gp63 gene locus (37). Indeed, a sequencing analysis could help to determine strain heterogeneity that link with geographical origin, also this technique detects genetic differentiation of clinical samples (38).

According to the results, the kDNA minicircle sequence of ten *L. tropica* from different geographical regions showed light heterogeneity. Phylogenetic tree showed a genetic affinity of all local *L. tropica* isolates were relatively more similar sequences and genetically closed related to the Iranian AB678350.1 strain, especially MN334662-MN334666 and MN334668 isolates. The result is harmonious with Al-Bajalan *et al* (39) were mentioned that phylogenetic analysis of *Leishmania* strains using the cytochrome b gene sequences in Iraqi borderline area showed closely related to the Iranian MRHO/IR/75/ER strain. One way or another, the reason may have related to the vector came from Iranian border. In addition, there are a large number of Iranian visiting to sacred places in central and south of Iraq.

## Conclusion

Nested PCR is suitable for *Leishmania* identification at the species level. This province is considered one of the foci of leishmaniasis, especially, *L. tropica*. The phylogenetic data of *L. tropica* in Thi-Qar needs more studies about molecular epidemiology. The phylogenetic tree may give us an idea about the extent of genetic convergence between local isolates and Iranian strains. Moreover, the similarity in kDNA gene nucleotide sequences of local isolates increase whenever approached from the Iranian border.

The genetic variation of kDNA minicircle sequences among *L. tropica* isolates may cause different clinical manifestations of the dermal lesion. Therefore, genetic heterogeneity diagnosis is important for detection of therapy, control and epidemiological studies when the features of *L. tropica* are clinically relevant.

## Data Availability

All data were found

## Acknowledgements

This study is part of PhD thesis for Mohammed H. Flaih. The authors would like to thank the public health department/Thi-Qar health center for their kind assistance, especially, Dr. Haider Hantosh of head of the center. Also, thanks to staff of dermatology in Hospitals for their helping during the sample collection. This study did not support financially.

